# Associations between social network characteristics and COVID-19 vaccination intention – the SaNAE study

**DOI:** 10.1101/2024.07.08.24309958

**Authors:** Lisanne CJ Steijvers, Céline JA van Bilsen, Stephanie Brinkhues, Sarah E Stutterheim, Rik Crutzen, Robert AC Ruiter, Christian JPA Hoebe, Nicole HTM Dukers-Muijrers

## Abstract

**Background:** Social networks, our social relationships, play a role in the spread of infectious diseases but also in infection prevention behaviors such as vaccination. Here, we aimed to assess which individual, interpersonal (social network characteristics), community and societal factors are associated with COVID-19 vaccination intention during the second wave of the COVID-19 pandemic.

**Methods:** The cross-sectional study utilized online questionnaire data collected between August and November 2020 in community-dwelling adults aged 40 years and older. COVID-19 vaccination intention was measured by assessing whether respondents were willing to receive a COVID-19 vaccination if the vaccines became available. At the time of data collection, vaccines were still in development. Associations between individual (sociodemographic variables, health, health concerns), interpersonal (social network characteristics including structure, function, and quality), community (social and labor participation) and societal factors (degree of urbanization), and the outcome variables COVID-19 vaccination intention (yes vs no, yes vs unsure, unsure vs no) were assessed in stepwise multivariable regression analyses.

**Results:** Of all participants (N=3,396), 59% reported a positive intention to vaccinate against COVID-19, 35% were unsure, and 6% had no intention. Men, individuals of older age, those with a college or university degree, and those concerned about their personal and family health were more likely to have the intention to vaccinate. Interpersonal factors associated included having a larger network size (social network structure) and a larger proportion of informational and emotional supporters (social network function). Living outside of urban areas, a societal factor was also associated with the intention to vaccinate.

**Conclusion:** In this study, we determined key characteristics of COVID-19 vaccination intention. Health promotion and vaccination communication strategies should focus not only on individual factors but also incorporate the social environment. Our findings highlight the importance of organizing social networks to mobilize social support for pandemic preparedness.

## INTRODUCTION

Infection prevention measures, such as washing hands frequently, wearing facemasks, or social distancing, as applied during the COVID-19 pandemic, are crucial to containing the spread of the virus.[1] Immunization is another effective public health strategy. Vaccines prevent and control infectious disease outbreaks [[2–4]. After sequencing the SARS-CoV-2 genome, rapid vaccine development started in early 2020, leading to approved vaccines by the end of the year.[5] However, vaccine effectiveness depends on uptake and individuals’ willingness to be vaccinated. This behavior is influenced by individual and social environmental factors, including interpersonal, community, and societal factors, as outlined in the socio-ecological framework. [6–9]. Key individual factors affecting COVID-19 vaccination behavior include gender, age, concerns about long-term effects, and risk perception defined by perceived susceptibility and perceived severity of disease [10,11].

The social environment includes communities and social networks, our social relationships. Social network members may influence each other’s vaccination intentions and behaviors. A study in the United States demonstrated that individuals with family members and friends who did not support COVID-19 vaccinations were also less likely to be vaccinated against COVID-19.[12] Moreover, people who had no intention to vaccinate against COVID-19 were also less likely to be surrounded by network members who were performing or supporting preventive measures.[13] Social support from network members is important in stimulating vaccination intention and uptake. Perceived social support from others is associated with a higher intention to vaccinate against COVID-19.[14] Previous studies have shown that social support was also positively associated with the uptake of other vaccinations such as influenza or pneumococcal vaccines [15,16].

Most studies examining factors associated with COVID-19 vaccination intention have focused on either individual factors or (social) environmental factors such as social support or trust in governmental institutions [13,17–21]. However, social environmental factors, including social networks, encompass more than merely social support. Social networks can be described based on their structure, function, and quality [17–19]. Structural social network characteristics include network size, i.e., the number of social relationships, and network diversity, which pertains to the variety of relationships (e.g., family, friends, neighbors, and colleagues). Network density indicates the interconnectedness between different types of relationships. Other structural aspects involve homogeneity of the network members in terms of age and gender, geographical proximity, and mode or frequency of contact. Functional social network characteristics encompass various forms of social support, including informational (advice), emotional (discussing important topics), or practical (help with jobs in or around the house) support.[17,19] Additionally, social strain, i.e., relationships perceived as burdensome, demanding, or involving criticism, along with the varying strengths of relationships (from strong to strained) can serve as proxies for assessing the quality of social networks.[19,20]

To the best of our knowledge, there is limited insight into the associations between COVID-19 vaccination intention and social environment factors including social network structure, function, and quality. In this study, we aimed to assess which social environmental (interpersonal, community, and societal) factors are associated with COVID-19 vaccination intention in independently living adults aged 40 years and older, in combination with individual factors. Gaining insight into the factors associated with vaccination intention helps better prepare for and respond to pandemics. It also highlights the importance of including the social environment in infection prevention strategies for pandemic preparedness.

## METHODS

### Study design and population

The research proposal of the current study was pre-registered [21] and relevant supportive materials are open access available. This cross-sectional study used data from the Dutch SaNAE cohort (www.sanae-study.nl) and was reported according to the STROBE guidelines.[22] The SaNAE cohort includes community-dwelling adults aged 40 years and older living in Limburg, the Netherlands. Between August and November 2020, 5,001 people were invited to complete an online questionnaire, 3,505 (67%) of whom completed the questionnaire. Respondents were slightly older (mean difference 2.2 years, *p*<.001), and more likely to have obtained a college or university degree (χ^2^ = 25,117; df=2; *p*<.001), but did not differ from non-responders in gender (χ^2^ =0,726; df=1; *p*=.394) or network size (mean difference −0.4, *p*=.112). In total, complete data were available for 3,396 participants.

### Measurements

#### COVID-19 vaccination intention

During data collection between August and November 2020, COVID-19 vaccines were not yet available, as they were still being developed, and approval from the European Medicines Agency (EMA) was pending.[23] Hence, intention was measured by asking participants if they were planning on getting the COVID-19 vaccine if it became available. Answer categories were yes, I don’t know (yet), and no.

#### Individual factors

Individual factors included were sociodemographic characteristics, health, and health concerns (Figure 1).

**Figure 1.**
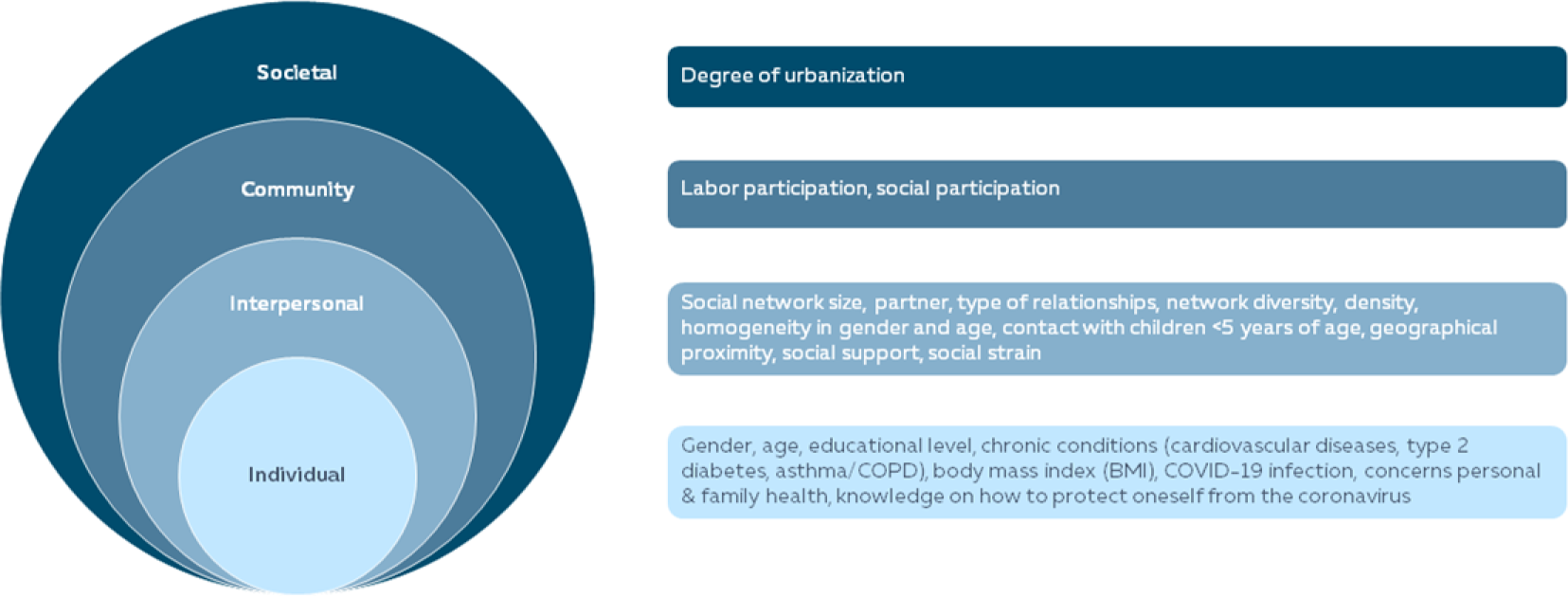
Overview of determinants of COVID-19 vaccination intention in the socio-ecological framework

##### Sociodemographic characteristics

Sociodemographic variables included gender (men/women/other) and age. Educational attainment was categorized into three levels: less than vocational education (no education, primary education (not completed), and lower vocational education), vocational education (intermediate vocational and higher secondary education), and college or university degree (higher professional and university education).[24]

##### Health

Health was determined by participants reporting any chronic conditions such as cardiovascular diseases, type 2 diabetes mellitus, or asthma/COPD. Body Mass Index (BMI) was calculated by dividing weight by height squared. Suspected SARS-CoV-2 infection was assessed by asking if they (think they) had been infected with the coronavirus. Answer categories were yes, maybe, and no.

##### Health concerns

Several health concerns during the COVID-19 pandemic were assessed. Concerns about one’s personal or family’s health were assessed on a five-point Likert scale with answer categories being very unconcerned, unconcerned, neither concerned nor unconcerned, concerned, or very concerned. Knowledge of how to protect oneself from the coronavirus was assessed using a five-point Likert scale with answer categories totally agree, agree, neutral, disagree, and totally disagree.

#### Interpersonal factors

Social networks were measured using a name-generator questionnaire.[25–27] A more detailed description of the name generator questionnaire with name interpreter items is available elsewhere. [28–30] Participants were asked to provide the names of up to fifteen family members, ten friends, ten acquaintances, five other persons, and five healthcare professionals (HCP) who are important to them or provide social support, up to a maximum of 45 network members. Social network size was then calculated by tallying all listed network members. Social network size was further categorized based on quartiles and used as an indicator for social isolation (category 0-4 network members). Additional information about the network members was collected using name interpreter items which were then used to describe social network structure, function, and quality, as described below (see also Supplementary Table 1).

##### Social network structure

Partner status was determined by a yes/no question. Relationship types were evaluated by calculating the proportion of each type (family, friends, acquaintances, others, HCP) in the total network. Network diversity was categorized based on the presence of combinations of different relationship types. Network density was measured by how well friends and family know each other, rated on a five-point scale. The homogeneity of social networks was assessed by the proportion of members of the same gender and age. Contact with children younger than five years of age was categorized by frequency (yes, daily or weekly; yes, monthly or less often; no), and living situation was determined by a single question, identifying those living alone. Geographical proximity was calculated by the proportion of network members living in the same house, within walking distance, less than thirty minutes away by car, more than thirty minutes away by car, or further away (Supplementary Table 1).[25–27]

##### Social network function

The proportion of network members who provided informational support was calculated by dividing the total number of network members providing informational support by the total network size. The proportion of network members who provide emotional support was calculated by dividing the total number of network members providing emotional support by the total network size. Lastly, the proportion of network members who provide practical support was calculated similarly.[25–27]

##### Social network quality

The proportion of network members with whom there is social strain was calculated by dividing the sum of the number of network members who are demanding, straining, or criticizing by the total network size. The proportion of network members with whom the relationship is good was calculated by dividing the number of network members with whom the relationship is good by the total network size. [25–27]

#### Community factors

##### Labor participation

Labor participation was assessed by asking participants if they were employed, (e.g., contract workers, freelancers), unemployed (due to incapacity, students or homemakers), or retired.

##### Social participation

Participation in social activities was evaluated by asking participants to report club memberships. This included sports clubs (e.g., sports or walking clubs), cultural organizations (music, dance, theater, carnival organizations), volunteer work, and other memberships.

#### Societal factors

##### Degree of urbanization

The 4-digit postal codes were converted into the degree of urbanization based on address density: rural areas (<500 addresses per km^2^), hardly urbanized areas (500 to 1000 addresses per km^2^), moderately urbanized areas (1000-1500 addresses per km^2^), strongly or extremely urbanized areas (>1500 addresses per km^2^).[31,32]

### Statistical analyses

Descriptive analyses were performed to describe the study population. Univariable logistic regression analyses were conducted with dummy variables of COVID-19 vaccination intention as the outcome variables (yes versus no, yes versus unsure, and unsure versus no), and all individual, interpersonal, community, and societal separately as independent categorical variables (except for age, BMI, network size, and proportions concerning network members, which were included as continuous variables). Correlations between variables were assessed and no multicollinearity was observed (all correlations < 0.7, VIF<10, and tolerance >0.1). All factors were then added stepwise per level in a multivariable logistic regression model using forward selection. Individual determinants were added in the first block, as these individual factors are most proximal to intention. Then, interpersonal, community, and societal factors were included to assess associations on all different levels. A *p*-value <.05 indicated statistical significance and all analyses were performed using IBM SPSS Statistics (version 27.0).

## RESULTS

### Study population

Among the participants, 55% were men and the mean age was 67 years. 59% of the participants reported intention to vaccinate against COVID-19, 35% were unsure, and 6% had no intention to vaccinate against COVID-19 (Table 1).

**Table 1.** Characteristics of the SaNAE study population in 2020 (n=3,396)

|  | COVID-19 vaccination intention |  |  |  |
| --- | --- | --- | --- | --- |
|  | Yes | Unsure | No |  |
|  | 59% (2001/3396) | 35% (1175/3396) | 6% (220/3396) |  |
| **Individual factors** |  |  |  |  |
| **Background characteristics** |  |  |  |  |
| Gender |  |  |  | *p*<.001 |
| Men | 63 (1259/2001) | 46 (536/1175) | 36 (79/220) |  |
| Women | 37 (742/2001) | 54 (639/1175) | 64 (141/220) |  |
| Age | 67.3 (9.4) | 62.6 (9.6) | 59.6 (9.1) | *p*<.001 |
| Educational attainment |  |  |  | *p*<.001 |
| Less than vocational training | 26 (529/2001) | 28 (323/1175) | 29 (63/220) |  |
| Vocational training | 25 (504/2001) | 35 (412/1175) | 42 (93/220) |  |
| College or university degree | 48 (968/2001) | 37 (440/1175) | 29 (64/220) |  |
| **Health** |  |  |  |  |
| Chronic conditions <sup>a</sup> | 32 (643/2001) | 25 (290/1175) | 25 (54/220) | *p*<.001 |
| BMI | 26.6 (4.4) | 27.0 (4.9) | 26.9 (4.5) | *p*=.101 |
| (suspected) COVID-19 infection |  |  |  | *p*=.002 |
| Yes/maybe | 18 (359/2001) | 22 (263/1175) | 25 (54/220) |  |
| No | 82 (1642/2001) | 78 (166/1175) | 75 (166/220) |  |
| **Health concerns** |  |  |  |  |
| Concerns about personal health |  |  |  | *p*<.001 |
| (very) Concerned | 26 (515/2001) | 18 (210/1175) | 11 (25/220) |  |
| Neutral | 36 (713/2001) | 40 (465/1175) | 30 (66/220) |  |
| (very) Concerned | 39 (773/2001) | 43 (500/1175) | 59 (129/220) |  |
| Concerns about family health |  |  |  | *p*<.001 |
| (very) Concerned | 41 (804/2001) | 35 (402/1175) | 27 (60/220) |  |
| Neutral | 36 (712/2001) | 39 (457/1175) | 32 (70/220) |  |
| (very) Unconcerned | 24 (485/2001) | 27 (316/1175) | 41 (90/220) |  |
| Knowledge of how to protect oneself from the COVID-19 virus |  |  |  | *p*<.001 |
| (totally) Agree | 87 (1731/2001) | 77 (909/1175) | 76 (167/220) |  |
| Neutral | 11 (223/2001) | 20 (238/1175) | 21 (47/220) |  |
| (totally) Disagree | 2 (47/2001) | 3 (28/1175) | 3 (6/220) |  |
| **Interpersonal factors** |  |  |  |  |
| **Social network structure** |  |  |  |  |
| Network size (including health care professionals) | 10.3 (7.3) | 10.5 (7.3) | 9.7 (7.0) | *p*=.339 |
| 0-4 network members | 25 (507/2001) | 25 (290/1175) | 26 (58/220) | *p*=.387 |
| 5-8 network members | 26 (514/2001) | 24 (276/1175) | 29 (64/220) |  |
| 9-12 network members | 20 (402/2001) | 22 (253/1175) | 16 (36/220) |  |
| ≥13 network members | 29 (578/2001) | 30 (356/1175) | 28 (62/220) |  |
| Partner | | | | $p=.079$ |
| Yes | 80 (1604/2001) | 78 (921/1175) | 74 (163/220) |  |
| No | 20 (397/2001) | 22 (254/1175) | 26 (57/220) |  |
| Type of relationships (proportion of network members:) |  |  |  |  |
| Family members | 51.1 (24.9) | 51.2 (25.1) | 51.9 (27.0) | $p=.907$ |
| Friends | 24.0 (18.2) | 24.9 (18.3) | 26.2 (21.3) | $p=.167$ |
| Acquaintances | 16.0 (15.5) | 16.1 (15.3) | 14.4 (14.6) | $p=.294$ |
| Extra members | 4.8 (11.5) | 4.6 (10.4) | 4.7 (10.5) | $p=.853$ |
| Health care professionals | 4.2 (9.4) | 3.3 (8.5) | 2.9 (7.9) | $p=.011$ |
| Network diversity | | | | $p=.248$ |
| No family, but friends, acquaintances, others, or HCP | 2 (38/2001) | 2 (19/1175) | 3 (6/220) |  |
| Only family, no friends/acquaintances/others/HCP | 12 (241/2001) | 13 (147/1175) | 15 (33/220) |  |
| Family and friends, no acquaintances/others/HCP | 14 (270/2001) | 15 (178/1175) | 16 (35/220) |  |
| Family and friends and acquaintances/others/HCP | 62 (1230/2001) | 62 (730/1175) | 58 (127/220) |  |
| Family and acquaintances/others/HCP, no friends | 11 (222/2001) | 9 (101/1175) | 9 (19/220) |  |
| Network density (my friends know my family) | | | | $p=.851$ |
| Yes | 75 (1507/2001) | 75 (880/1175) | 74 (162/220) |  |
| No | 25 (494/2001) | 25 (295/1175) | 26 (58/220) |  |
| Proportion of network members of the same gender | 52.1 (22.6) | 55.8 (23.9) | 55.2 (22.7) | $p<.001$ |
| Proportion of network members of the same age | 39.4 (23.9) | 39.9 (23.8) | 38.4 (24.1) | $p=.674$ |
| Contact with children <5 years of age | | | | $p=.002$ |
| Yes, daily or weekly | 20 (405/2001) | 22 (261/1175) | 27 (59/220) |  |
| Yes, monthly or less often | 20 (392/2001) | 24 (279/1175) | 18 (40/220) |  |
| No | 60 (1204/2001) | 54 (635/1175) | 55 (121/220) |  |
| Living alone | 21 (413/2001) | 21 (241/1175) | 25 (56/220) | $p=.229$ |
| Geographical proximity (proportion of network members who live:) |  |  |  |  |
| In the same house | 15.8 (20.7) | 16.3 (21.2) | 18.6 (24.1) | $p=.173$ |
| Within walking distance | 29.0 (25.5) | 27.5 (24.4) | 28.1 (26.1) | $p=.267$ |
| Less than 30 minutes away | 35.7 (27.0) | 38.4 (26.4) | 38.7 (28.5) | $p=.014$ |
| More than 30 minutes away | 10.2 (16.3) | 9.8 (15.9) | 8.3 (14.1) | $p=.234$ |
| Further away | 9.1 (16.1) | 7.8 (14.8) | 6.0 (14.6) | $p=.005$ |
| **Social network function** |  |  |  |  |
| Proportion of network members who provide informational support | 49.3 (33.4) | 50.6 (33.5) | 46.9 (34.2) | $p=.279$ |
| Proportion of network members who provide emotional support | 63.4 (31.2) | 61.4 (31.4) | 62.8 (31.6) | $p=.195$ |
| Proportion of network members who provide practical support | 22.3 (24.9) | 21.5 (23.9) | 21.5 (23.0) | $p=.669$ |
| **Social network quality** |  |  |  |  |
| Proportion of network members with whom the relationship is demanding, straining, or criticizing (social strain) | 9.9 (18.5) | 11.3 (19.5) | 9.8 (15.7) | $p=.102$ |
| Proportion of network members with whom there is a good relationship: | 94.9 (23.0) | 93.2 (17.6) | 92.7 (14.0) | $p=.038$ |
| **Community factors** |  |  |  |  |
| Labor participation |  |  |  |  |
| Employed | 26 (515/2001) | 42 (498/1175) | 53 (117/220) | $p<.001$ |
| Retired | 52 (1034/2001) | 34 (398/1175) | 21 (47/220) |  |
| Unemployed | 23 (452/2001) | 24 (279/1175) | 26 (56/220) |  |
| Social participation (club membership) |  |  |  |  |
| Sports club membership | 33 (625/2001) | 31 (369/1175) | 28 (61/220) | $p=.313$ |
| Music organization membership | 15 (306/2001) | 12 (139/1175) | 12 (27/220) | $p=.019$ |
| Volunteer work | 16 (323/2001) | 13 (154/1175) | 10 (22/220) | $p=.008$ |
| Other (religious groups, talking groups, etc.) | 25 (509/2001) | 22 (259/1175) | 21 (45/220) | $p=.044$ |
| **Societal factors** |  |  |  |  |
| Degree of urbanization | | | | $p<.001$ |
| Rural areas | 28 (554/2001) | 28 (334/1175) | 19 (42/220) |  |
| Hardly urbanized areas | 25 (493/2001) | 23 (270/1175) | 21 (47/220) |  |
| Moderately urbanized areas | 19 (372/2001) | 17 (204/1175) | 26 (57/220) |  |
| (Extremely) strongly urbanized areas | 29 (582/2001) | 31 (367/1175) | 34 (74/220) |  |

Numbers are presented in % (n/N) or mean (sd)
<sup>a</sup> chronic conditions include self-reported cardiovascular diseases, type 2 diabetes mellitus, asthma, and COPD.
<sup>#</sup> $p<.100$ , <sup>\*</sup> $p<.05$ , <sup>\*\*</sup> $p<.01$ , <sup>\*\*\*</sup> $p<.001$

Among participants with the intention to vaccinate against COVID-19, 26% were concerned about personal health, 41% were concerned about family health, and 87% indicated knowing what to do to protect themselves. Among participants who were unsure or had no intention to get vaccinated, 77% and 76% respectively, indicated knowing what to do to protect themselves.

### Overall social network characteristics

On average, participants, regardless of intention to vaccinate, had ten network members. Of these, slightly more than half were family members, a quarter were friends, and the remainder were acquaintances, neighbors, colleagues, informal healthcare professionals, or others (Table 1). Participants reported receiving informational support (information or advice) from half of their network members and emotional support (discussing important topics or health status) from just over 60% of their network members.

### Determinants of COVID-19 vaccination intention

#### COVID-19 vaccination intention: yes versus no

Individual factors associated with COVID-19 vaccination intention versus no intention were gender (men), older age, having obtained a college or university degree, having chronic conditions, having no previous (suspected) COVID-19 infection, having concerns about one’s personal and family health, and knowledge of how to protect oneself from the virus (Table 2). Interpersonal factors associated with COVID-19 vaccination intention were having a partner and having a larger proportion of network members who live far away. Community factors associated with COVID-19 intention were being unemployed or retired (versus employed) and doing volunteer work. Living in rural areas was a societal factor associated with COVID-19 intention (Table 2).

**Table 2.** Univariable logistic regression analyses of individual, interpersonal, community, and societal factors and COVID-19 vaccination intention.

|  | COVID-19 vaccination intention |  |  |
| --- | --- | --- | --- |
|  | Yes versus no | Yes versus unsure | Unsure versus no |
|  | OR (95% CI) | OR (95% CI) | OR (95% CI) |
| **Individual factors** |  |  |  |

| Background characteristics |  |  |  |
| --- | --- | --- | --- |
| Gender |  |  |  |
| Men | 3.03 (2.27-4.05)*** | 2.02 (1.75-2.34)*** | 1.50 (1.11-2.02)** |
| Women | Ref | Ref | Ref |
| Age | 1.08 (1.07-1.10)*** | 1.05 (1.04-1.06)*** | 1.03 (1.02-1.05)*** |
| Educational attainment |  |  |  |
| Less than vocational training | Ref | Ref | Ref |
| Vocational training | 0.65 (0.46-0.91)* | 0.75 (0.62-0.90)** | 0.86 (0.61-1.23) |
| College or university degree | 1.80 (1.25-2.59)** | 1.34 (1.12-1.61)** | 1.34 (0.92-1.95) |
| Health |  |  |  |
| Chronic conditions <sup>a</sup> | 1.46 (1.06-2.01)* | 1.45 (1.23-1.70)*** | 1.01 (0.72-1.41) |
| BMI | 0.99 (0.96-1.02) | 0.98 (0.97-1.00)* | 1.00 (0.97-1.04) |
| (suspected) COVID-19 infection |  |  |  |
| Yes/maybe | Ref | Ref | Ref |
| No | 1.49 (1.07-2.07)* | 1.32 (1.10-1.58)** | 1.13 (0.81-1.58) |
| Health concerns |  |  |  |
| Concerns about personal health |  |  |  |
| (very) Concerned | 3.44 (2.21-5.35)*** | 1.59 (1.30-1.93)*** | 2.17 (1.37-3.43)*** |
| Neutral | 1.80 (1.32-2.47)*** | 0.99 (0.84-1.17) | 1.82 (1.32-2.51)*** |
| (very) Unconcerned | Ref | Ref | Ref |
| Concerns about family health |  |  |  |
| (very) Concerned | 2.49 (1.76-3.51)*** | 1.30 (1.08-1.57)** | 1.91 (1.33-2.73)*** |
| Neutral | 1.89 (1.35-2.63)*** | 1.02 (0.84-1.22) | 1.86 (1.32-2.62)*** |
| (very) Unconcerned | Ref | Ref | Ref |
| Knowledge of how to protect oneself from the COVID-19 virus |  |  |  |
| (totally) Agree | 2.19 (1.54-3.11)*** | 2.03 (1.67-2.48)*** | 1.08 (0.76-1.53) |
| Neutral | Ref | Ref | Ref |
| (totally) Disagree | 1.65 (0.67-4.09) | 1.79 (1.08-2.96)* | 0.92 (0.36-2.35) |
| Interpersonal factors <sup>b</sup> |  |  |  |
| Social network structure |  |  |  |
| Network size (including health care professionals) | 1.01 (0.99-1.03) | 1.00 (0.99-1.01) | 1.02 (1.00-1.04) |
| Partner |  |  |  |
| Yes | 1.41 (1.03-1.95)* | 1.11 (0.93-1.33) | 1.27 (0.91-1.77) |
| No | Ref | Ref | Ref |
| Type of relationships (proportion of network members:) |  |  |  |
| Family members | 1.00 (0.56-1.79) | 0.95 (0.69-1.29) | 1.07 (0.58-1.96) |
| Friends | 0.50 (0.24-1.02)<sup>#</sup> | 0.79 (0.53-1.19) | 0.63 (0.30-1.33) |
| Acquaintances | 1.75 (0.66-4.67) | 0.97 (0.59-1.58) | 1.80 (0.64-5.10) |
| Extra members | 1.02 (0.30-3.42) | 1.24 (0.64-2.40) | 0.77 (0.20-2.97) |
| Health care professionals | 5.84 (0.96-35.47)<sup>#</sup> | 2.98 (1.29-6.87)* | 1.86 (0.31-11.19) |
| Network diversity |  |  |  |
| No family, but friends, acquaintances, others, or HCP | 0.87 (0.34-2.21) | 1.22 (0.68-2.20) | 0.71 (0.26-1.92) |
| Only family, no friends/acquaintances/others/HCP | Ref | Ref | Ref |
| Family and friends, no acquaintances/others/HCP | 1.06 (0.64-1.75) | 0.93 (0.70-1.22) | 1.14 (0.68-1.93) |
| Family and friends and acquaintances/others/HCP | 1.33 (0.88-1.99) | 1.03 (0.82-1.29) | 1.29 (0.85-1.97) |
| Family and acquaintances/others/HCP, no friends | 1.60 (0.88-2.90) | 1.34 (0.98-1.83) <sup>#</sup> | 1.19 (0.64-2.22) |
| Network density (my friends know my family) |  |  |  |
| Yes | 1.09 (0.80-1.50) | 1.02 (0.87-1.21) | 1.07 (0.77-1.48) |
| No | Ref | Ref | Ref |
| Proportion of network members with different gender | 1.88 (0.99-3.57)<sup>#</sup> | **2.02 (1.47-2.77)\*\*\*** | 0.90 (0.49-1.66) |
| Proportion of network members of the same age | 1.19 (0.67-2.12) | 0.92 (0.68-1.25) | 1.27 (0.70-2.30) |
| Contact with children <5 years of age |  |  |  |
| Yes, daily or weekly | Ref | Ref | Ref |
| Yes, monthly or less often | 1.43 (0.93-2.18) | 0.91 (0.73-1.13) | 1.19 (0.84-1.67) |
| No | **1.45 (1.04-2.02)\*** | **1.22 (1.02-1.47)\*** | **1.58 (1.02-2.44)\*** |
| Living alone | 0.76 (0.55-1.05)<sup>#</sup> | 1.01 (0.84-1.21) | 0.76 (0.54-1.06) |
| Geographical proximity (proportion of network members who live:) |  |  |  |
| In the same house | 0.61 (0.31-1.19) | 0.83 (0.57-1.20) | 0.74 (0.37-1.46) |
| Within walking distance | 1.11 (0.64-1.92) | 1.28 (0.96-1.72)<sup>#</sup> | 0.88 (0.49-1.56) |
| Less than 30 minutes away | 0.65 (0.39-1.07) | **0.69 (0.53-0.91)\*\*** | 0.89 (0.52-1.53) |
| More than 30 minutes away | 2.10 (0.81-5.48) | 1.18 (0.75-1.86) | 1.81 (0.67-4.87) |
| Further away | **3.86 (1.33-11.21)\*** | **1.74 (1.09-2.79)\*** | 2.18 (0.75-6.40) |
| **Social network function** |  |  |  |
| Proportion of network members who provide informational support | 1.28 (0.84-1.94) | 0.89 (0.72-1.10) | 1.42 (0.93-2.18) |
| Proportion of network members who provide emotional support | 1.09 (0.70-1.70) | 1.23 (0.98-1.55) <sup>#</sup> | 0.89 (0.56-1.40) |
| Proportion of network members who provide practical support | 1.24 (0.69-2.22) | 1.12 (0.82-1.51) | 1.12 (0.61-2.09) |
| **Social network quality** |  |  |  |
| Proportion of network members with whom the relationship is demanding, straining, or criticizing (social strain) | 0.98 (0.45-2.10) | **1.47 (1.01-2.14)\*** | 0.64 (0.28-1.45) |
| Proportion of network members with whom there is a good relationship: | 2.25 (0.95-5.33)<sup>#</sup> | **1.76 (1.11-2.78)\*** | 1.14 (0.50-2.62) |
| **Community factors** |  |  |  |
| Labor participation |  |  |  |
| Employed | Ref | Ref | Ref |
| Retired | **5.00 (3.51-7.13)\*\*\*** | **2.51 (2.12-2.98)\*\*\*** | **1.99 (1.38-2.86)\*\*\*** |
| Unemployed | **1.83 (1.30-2.58)\*\*\*** | **1.57 (1.29-1.90)\*\*\*** | 1.17 (0.82-1.66) |
| Social participation (club membership) |  |  |  |
| Sports club membership | 1.26 (0.92-1.72) | 1.06 (0.90-1.23) | 1.19 (0.87-1.64) |
| Music organization membership | 1.29 (0.85-1.97) | **1.35 (1.09-1.67)\*\*** | 0.96 (0.62-1.49) |
| Volunteer work | **1.73 (1.10-2.73)\*** | **1.28 (1.04-1.57)\*** | 1.36 (0.85-2.18) |
| Other (religious groups, talking groups, etc.) | 1.33 (0.94-1.87) | **1.21 (1.02-1.43)\*** | 1.10 (0.77-1.57) |
| **Societal factors** |  |  |  |
| Degree of urbanization |  |  |  |
| Rural areas | **1.68 (1.13-2.49)\*** | 1.05 (0.87-1.26) | **1.60 (1.07-2.41)\*** |
| Hardly urbanized areas | 1.33 (0.91-1.96) | 1.15 (0.95-1.40) | 1.16 (0.78-1.72) |
| Moderately urbanized areas | 0.83 (0.57-1.20) | 1.15 (0.93-1.43) | 0.72 (0.49-1.06)<sup>#</sup> |
| (Extremely) strongly urbanized areas | Ref | Ref | Ref |

<sup>a</sup> chronic conditions include self-reported cardiovascular diseases, type 2 diabetes mellitus, asthma, and COPD.
<sup>b</sup> all proportion variables are adjusted for network size
OR (95% CI) = Odds Ratio (95% Confidence Interval)
<sup>#</sup> $p < .100$ , \* $p < .05$ , \*\* $p < .01$ , \*\*\* $p < .001$

#### COVID-19 vaccination intention: yes versus unsure

Individual factors associated with COVID-19 vaccination intention versus being unsure were gender (men), older age, having obtained a college or university degree, having chronic conditions, having no previous (suspected) COVID-19 infection, being concerned about personal and family health and knowledge of how to protect oneself from the virus (Table 2). A higher BMI was inversely associated with COVID-19 vaccination intention. Interpersonal factors associated were having a larger proportion of network members who were healthcare professionals, a larger proportion of network members of a different gender, no contact with children younger than five years of age, a larger proportion of network members living further away, a larger proportion of network members with whom there is no social strain, and a larger proportion with whom the relationship is good. Community factors associated with COVID-19 intention were being retired or unemployed (versus employment), having a music organization membership, doing volunteer work, or having other club memberships.

#### COVID-19 vaccination intention: unsure vs no

Individual factors associated with being unsure about getting a COVID-19 vaccination versus no intention were gender (men), older age, and having concerns about one’s personal and family health (Table 2). Not having contact with children younger than five years of age was an interpersonal factor associated with being unsure about getting a vaccination. A community factor associated with being unsure was being retired (versus employed). Living in rural areas was a societal factor associated with being unsure about getting a COVID-19 vaccination.

#### Multivariable logistic regression models COVID-19 vaccination intention

After including all variables in the multivariable models, the individual factors gender, age, and concerns about family health remained significantly associated with COVID-19 vaccination intention compared to no intention (Table 3), intention compared to being unsure (Table 4), and being unsure compared to no intention (Table 5). Additionally, educational attainment, concerns about personal health, and knowledge of how to protect oneself from the virus remained associated with both intention versus no intention and intention versus being unsure. Interpersonal factors such as network size and proportion of informational supporters were associated with intention versus no intention, whereas the proportion of emotional supporters was associated with intention versus being unsure. Furthermore, club membership in a music organization, a community factor, was associated with intention versus being unsure. Lastly, at the societal level, living in rural areas was associated with intention or being unsure versus no intention.

**Table 3.**
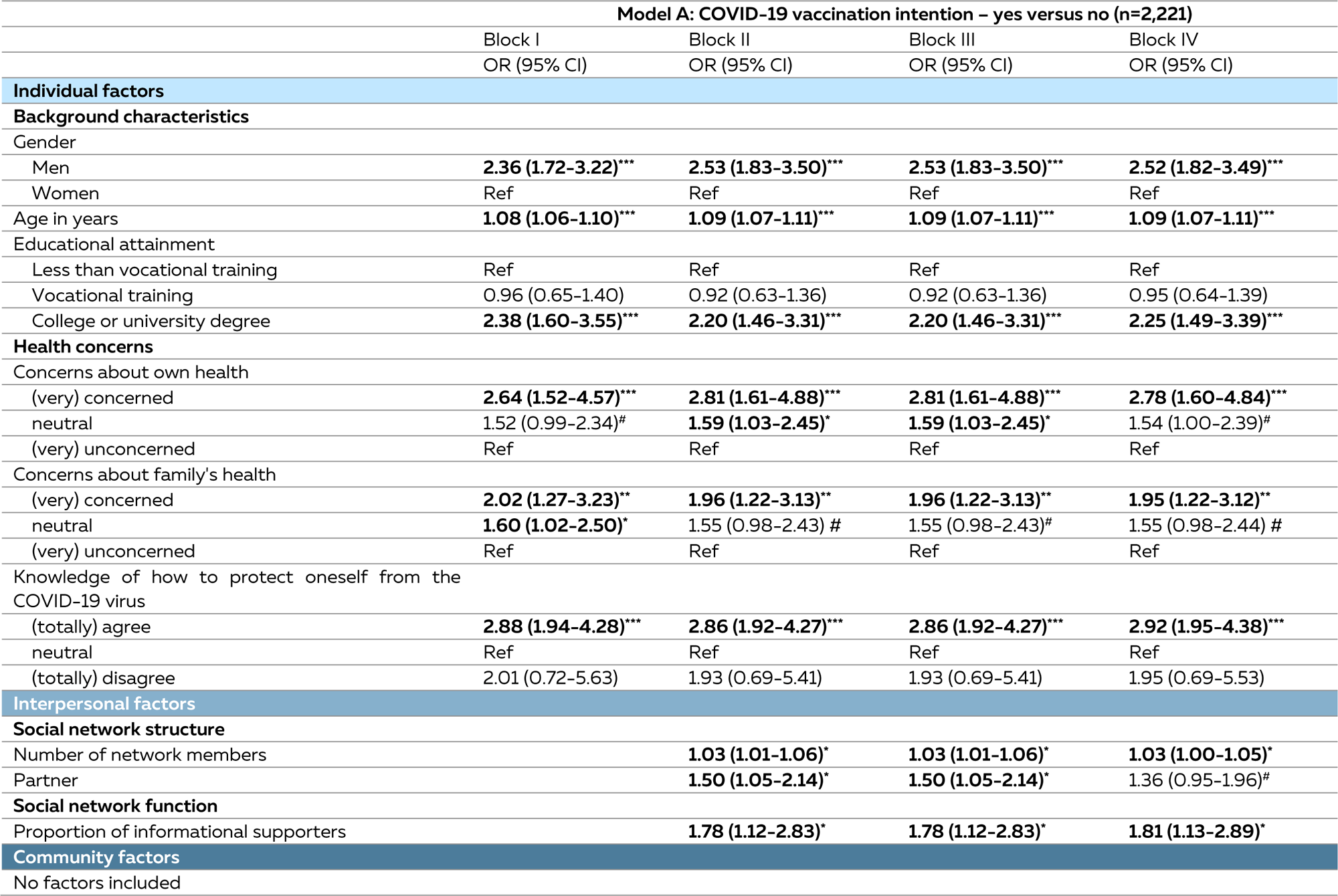
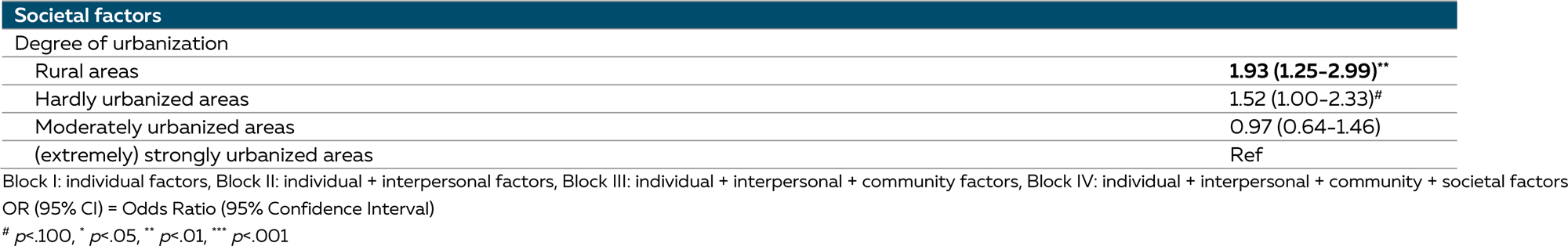
Multivariable logistic regression analyses of individual, interpersonal, community, and societal factors and COVID-19 intention (yes versus no)

**Table 4.**
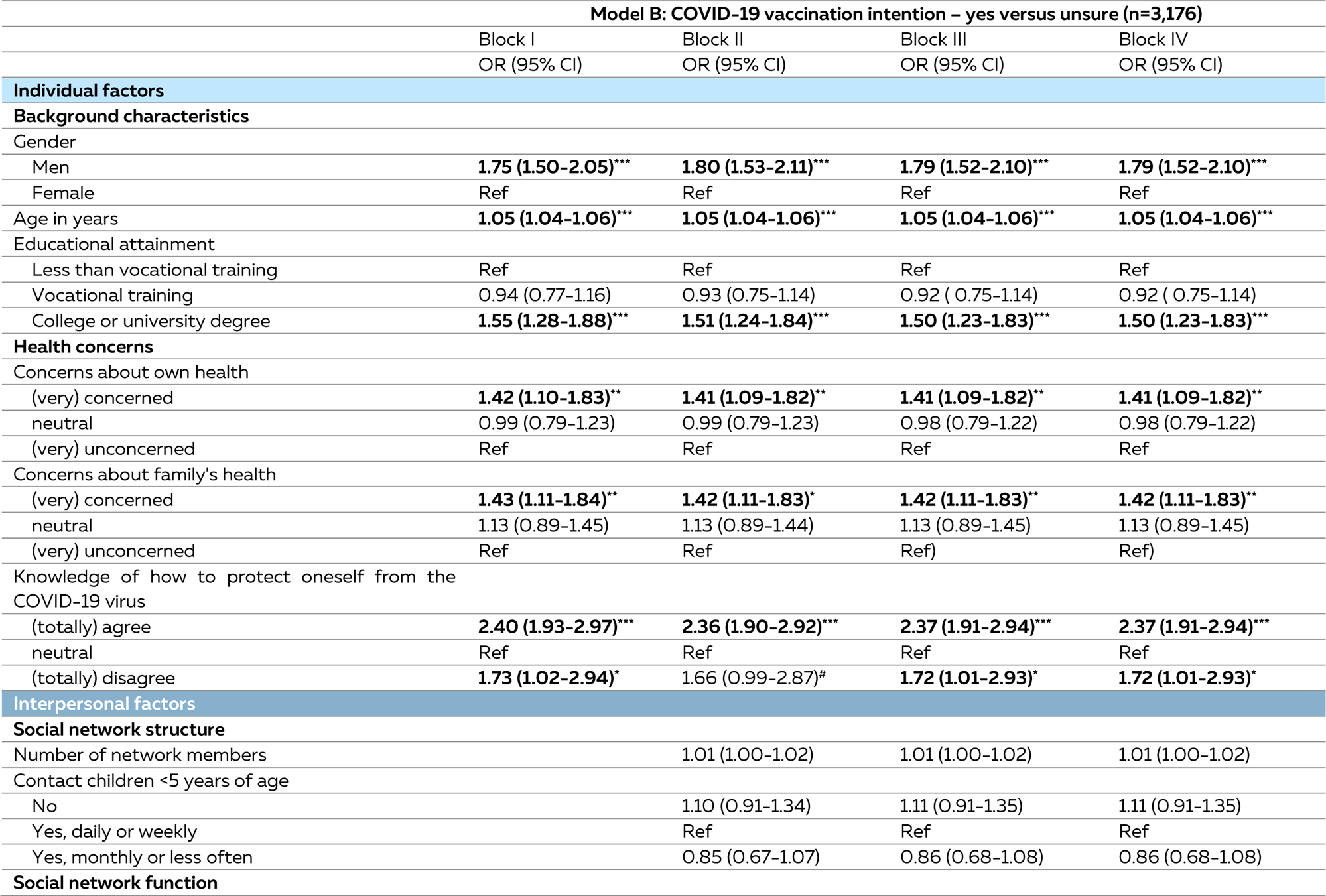
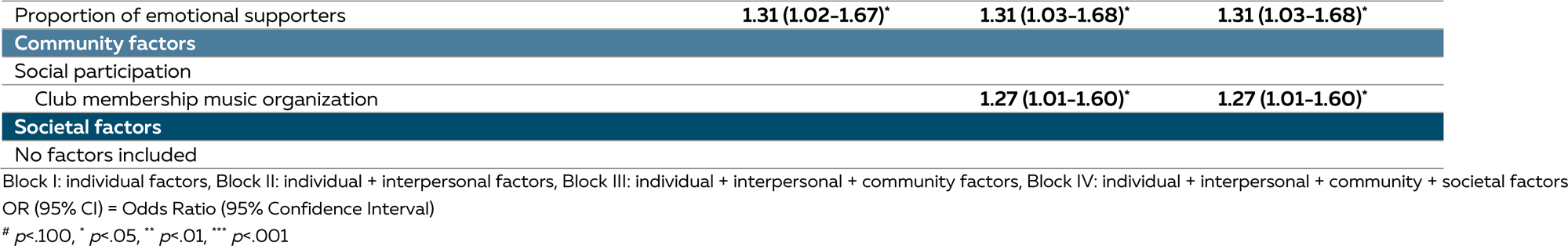
Multivariable logistic regression analyses of individual, interpersonal, community, and societal factors and COVID-19 intention (yes versus unsure)

**Table 5.** Multivariable logistic regression analyses of individual, interpersonal, community, and societal factors and COVID-19 intention (unsure vs no)

| Model C: COVID-19 vaccination intention – unsure versus no (n=1,395) |  |  |  |  |
| --- | --- | --- | --- | --- |
|  | Block I | Block II | Block III | Block IV |
|  | OR (95% CI) | OR (95% CI) | OR (95% CI) | OR (95% CI) |
| **Individual factors** |  |  |  |  |
| **Background characteristics** |  |  |  |  |
| Gender |  |  |  |  |
| Men | 1.42 (1.04-1.93)* | 1.52 (1.10-2.08)* | 1.52 (1.10-2.08)* | 1.58 (1.15-2.18)** |
| Women | Ref | Ref | Ref | Ref |
| Age in years | 1.04 (1.02-1.05)*** | 1.04 (1.02-1.05)*** | 1.04 (1.02-1.05)*** | 1.04 (1.02-1.05)*** |
| Educational attainment |  |  |  |  |
| Less than vocational training | Ref | Ref | Ref | Ref |
| Vocational training | 1.02 (0.71-1.48) | 0.98 (0.68-1.42) | 0.98 (0.68-1.42) | 0.97 (0.67-1.41) |
| College or university degree | 1.65 (1.12-2.45)* | 1.53 (1.02-2.28)* | 1.53 (1.02-2.28)* | 1.47 (0.98-2.20)# |
| **Health concerns** |  |  |  |  |
| Concerns about family's health |  |  |  |  |
| (very) concerned | 2.26 (1.56-3.27)*** | 2.25 (1.55-3.25)*** | 2.25 (1.55-3.25)*** | 2.36 (1.62-3.43)*** |
| neutral | 1.97 (1.39-2.80)*** | 1.99 (1.40-2.82)*** | 1.99 (1.40-2.82)*** | 1.96 (1.38-2.80)*** |
| (very) unconcerned | Ref | Ref | Ref | Ref |
| **Interpersonal factors** |  |  |  |  |
| **Social network structure** |  |  |  |  |
| Number of network members |  | 1.02 (1.00-1.04)# | 1.02 (1.00-1.04)# | 1.02 (1.00-1.04) |
| **Community factors** |  |  |  |  |
| No factors included |  |  |  |  |
| **Societal factors** |  |  |  |  |
| Degree of urbanization |  |  |  |  |
| Rural areas |  |  |  | 1.69 (1.11-2.57)* |
| Hardly urbanized areas |  |  |  | 1.17 (0.78-1.76) |
| Moderately urbanized areas |  |  |  | 0.73 (0.49-1.08) |
| (extremely) strongly urbanized areas |  |  |  | Ref |

Block I: individual factors, Block II: individual + interpersonal factors, Block III: individual + interpersonal + community factors, Block IV: individual + interpersonal + community + societal factors
OR (95% CI) = Odds Ratio (95% Confidence Interval)

## DISCUSSION

In this study, we assessed associations between individual, interpersonal, community, and societal factors and COVID-19 vaccination intention among adults aged 40 years and older living independently in the Netherlands. Of all participants, 59% had the intention to get a COVID-19 vaccination if it would become available, 35% were unsure, and 6% had no intention.

Individual factors associated with COVID-19 vaccination intention (yes versus no) included sociodemographic characteristics such as gender, age, and educational attainment. Men, older individuals, and those who had obtained a college or university degree were more likely to have the intention to get a COVID-19 vaccination. Gender and age were also associated with having intention versus being unsure and being unsure versus no intention. Additionally, educational attainment was associated with having the intention versus being unsure. These findings align with previous studies that identified sociodemographic characteristics as significant predictors of COVID-19 vaccination intention.[33–35] However, there are some inconsistencies regarding age. Some studies indicated that younger individuals were more likely to express willingness to receive a COVID-19 vaccination, which is in contrast to our findings. The majority of the studies reported higher willingness among older individuals.[36,37] The higher levels of intention observed among men and older individuals may be explained by their increased risk of contracting the coronavirus, experiencing more severe infections, and facing higher mortality rates.[38–40]

Moreover, concerns about personal and family health and knowledge of how to protect oneself from the coronavirus were associated with COVID-19 vaccination intention as opposed to being unsure. Previous studies have established that individuals worried about their health, those more vulnerable, and those at higher risk for COVID-19 infection were more willing to get vaccinated.[35] Additionally, concerns about family health, in terms of potential coronavirus infections, were also associated with being more willing to vaccinate against COVID-19.[41] These findings suggest that concerns about personal and family health contribute to higher vaccination intentions, highlighting the importance of achieving herd immunity to safeguard social network members [42]

In addition to individual factors, interpersonal, community, and societal factors such as social support, club memberships, and degree of urbanization, were also associated with COVID-19 vaccination intention. Individuals who had a larger network size and a larger proportion of network members who provide informational support were more likely to have the intention to vaccinate against COVID-19 (versus no intention), and those who had a larger proportion of emotional supporters within the network were also more likely to have the intention to vaccinate against COVID-19 (versus being unsure). Previous studies have already established that higher perceived social support is associated with willingness to vaccinate and higher uptake of the COVID-19 vaccination.[43,44] This might be explained by the theory of strong and weak ties by Granovetter[37], which postulated that weaker social ties (e.g., acquaintances or new network members) play an important role in the provision of information. However, in the current study, it remained unknown what kind of information was exchanged and whether this was related to vaccination intention. Higher levels of social support and increased social contact may also expose individuals to diverse views on COVID-19, helping them assess their own risk and need for the vaccine.[43] The social support roles of the network members emphasize the need to strengthen or expand social networks to have larger and more diverse networks in which different types of social support are provided, especially in times of a pandemic.

Lastly, participants living in rural areas versus those living in urban areas were more likely to have the intention or be unsure of getting a COVID-19 vaccination (versus no intention). These results are inconsistent with previous studies assessing COVID-19 vaccination intention and urban-rural differences. Several studies have reported that people living in rural areas are less willing to vaccinate or have lower actual vaccination rates.[46,47] An explanation for these contradicting results might be related to the network composition for middle-aged and older adults in the Southern part of the Netherlands. Individuals living in rural areas tended to have larger and more diverse social networks with more social supporters compared to those living in urban areas.[29] We suggest future research take a neighborhood-specific approach.

### Implications

The various factors identified in this study present opportunities for health promotion. These key characteristics associated with vaccination intention can be incorporated into a practical toolkit to inform researchers, healthcare professionals, and policymakers to identify sociodemographic groups with potentially low vaccination intentions, allowing for a focused approach. One example might be a neighborhood-specific approach to promote health behaviors such as vaccination. For instance, mobile vaccination buses were introduced during the COVID-19 pandemic to increase vaccine uptake in neighborhoods with low vaccination rates.[48] Additionally, offering vaccinations at more accessible locations within communities, such as pharmacies, could contribute to increasing vaccination intention and uptake.[49,50]

In addition to individual factors, interpersonal factors such as social network size, and informational, and emotional social support were evident, emphasizing the need to strengthen and expand social networks and mobilize social support roles within the network. Strengthening and expanding social networks is particularly relevant for pandemic preparedness, as large, diverse, and supportive social networks can act as buffers during stressful times, such as pandemics.[29,51] Moreover, networks are valuable at all times, contributing to overall health, well-being, and resilience.[19] While directly modifying social networks might be challenging, we argue that they might be influenced indirectly through policy interventions. For example, policies can address other environmental factors such as important local influencers in communities [48] or implementing changes to the physical environment, thereby enhancing the social environment and creating opportunities for social interaction.

### Strengths and limitations

The strengths of this study are the inclusion of multiple levels of the socio-ecological model. In doing so, social networks were measured using a name generator questionnaire which is a reliable method for a detailed assessment of social networks, especially in larger surveys.[25–27] With this method, a distinction in social network characteristics can be made, allowing for the inclusion of a broad range of social network aspects and differentiating between structure, function, and quality of social networks rather than just focusing on network size and social support. However, limitations should be mentioned as well. During the period of data collection, vaccines were still in development. Therefore, it was not possible to include actual vaccination uptake in this study. Future studies should investigate whether the individual, interpersonal, community, and societal factors identified in this study are also associated with actual COVID-19 vaccination uptake.

## CONCLUSION

In the present study, we aimed to assess which individual, interpersonal, community, and societal factors are associated with COVID-19 vaccination intention. Key individual determinants include various sociodemographic characteristics, concerns about one’s personal and family health, and knowledge of how to protect oneself from the virus. Beyond individual factors, informational and emotional social support at the interpersonal levels also plays a significant role. These findings suggest that health promotion and vaccination communication strategies should focus on these factors and highlight the importance of organizing social networks to mobilize social support, particularly during a pandemic.

## Data Availability

Data produced in the present study are available upon reasonable request to the authors

## Acknowledgments

The authors would like to thank Charlotte Anraad, Haiyue Shan, David Blanco-Herrero, Mirjam Kretzschmar, Danielle Timmermans, Bas van den Putte, and Vincent Buskens for their valuable contributions to this work.

## Funding

This work was funded by a governmental organization grant from the Dutch Organization for Health Research and Development (ZonMW Netherlands) (project number BePrepared: 10710022210002). The funder of the study had no role in the study design, data collection, data analysis, data interpretation, and writing of the manuscript.

## Ethics statement

The Medical Ethical Committee of Maastricht University approved this study (METC 2018-0698, 2019-1035, and 2020-2266). Participants gave electronic informed consent.

## Declaration of generative AI in scientific writing

During the preparation of this work, the author(s) used ChatGPT to improve readability and grammar. After using this tool/service, the author(s) reviewed and edited the content as needed and take(s) full responsibility for the content of the published article.

## SUPPLEMENTARY MATERIALS

**Supplementary Table 1.** Overview of social network characteristics described by structure, function, and quality.

| **Social network structure** |  |
| --- | --- |
| **Network size and social isolation** | Respondents could report up to fifteen family members, ten friends, ten acquaintances, five other network members, and five healthcare professionals (HCP). The sum of all network members per relationship type provided the total network size, resulting in a maximum of 45 persons. Network size was then further categorized based on quartiles: 0–4, 5–8, 9–13, and more than 13 network members. Network size was categorized to provide a quantitative measure for social isolation (0–4 network members) |
| **Partner** | Partner was assessed by a single question: do you have a partner? Answer categories included yes or no. |
| **Type of relationships** | The proportions of types of relationships were calculated by dividing the number of family members by the total network size. The same was performed for friends, acquaintances, other network members, and healthcare professionals. |
| **Network diversity** | Social network diversity was constructed based on the relationship types reported and included the following categories:<br>• Having no family, but friends, acquaintances, others, or HCP;<br>• Having only family members;<br>• Having only family members and friends;<br>• Having family members, others, HCP, but no friends<br>• Having family members, friends, acquaintances, other network members, and HCP. |
| **Network density** | Respondents could answer a 5-scaled statement about whether their friends and family know each other. |
| **Homogeneity in gender and age** | The proportion of network members of the same gender was calculated by dividing the number of network members of the same gender by the total network size. The proportion of network members of the same age (~5-year age range) was calculated by dividing the number of network members of the same age by the total network size. |
| **Contact with children aged five years and younger** | Contact with young children aged five years and younger was assessed by a single question with answer categories: yes daily, yes weekly, yes monthly, yes less often, no. Answer categories yes daily and yes weekly were combined into yes, daily, or weekly, and categories yes monthly and yes less often were combined into yes, monthly, or less often. |
| **Living alone** | The living situation was assessed by a single question with several answer categories: living alone, living with a partner, living with children, living with parents, living with other adults. Living alone was constructed based on the answer category: living alone. |
| **Geographical proximity** | Proportions of network members who live in the same house, within walking distance, less than 30 minutes away by car, more than 30 minutes away by car, or further away were calculated by dividing the number of network members living in the same house by the total network size, and so on. |
| **Social network function** |  |
| **Informational support** | The proportion of informational supporters was calculated by dividing the number of network members who advised on problems or gave information by the total network size. |
| **Emotional support** | The proportion of emotional supporters was calculated by dividing the number of network members who provided the opportunity to discuss important matters or health-related topics by the total network size. |
| **Practical support** | The proportion of practical supporters was calculated by dividing the number of network members who helped with small or larger tasks in or around the house by the total network size. |
| **Social network quality** |  |
| **Social strain** | The proportion of network members with whom social strain was experienced was calculated by dividing the sum of network members who are demanding, straining, or criticizing by the total network size. |
| **Good relationships** | The proportion of network members with whom there is a good relationship was calculated by dividing the number of social network members with whom there is a good relationship by the total network size. |

## Notes

### Competing Interest Statement

The authors have declared no competing interest.

### Author Declarations

The Medical Ethical Committee of Maastricht University approved this study (METC 2018-0698, 2019-1035, and 2020-2266).

